# Plasma Proteomic Profiles Predict the Risk of Coronary Artery Disease in the UK Biobank Cohort

**DOI:** 10.1101/2024.12.30.24319756

**Authors:** Chenhao Zhang, Xiangwen Ji, Chunmei Cui, Xueke Bai, Guangda He, Liang Chen, Qinghua Cui

**Affiliations:** Department of Biomedical Informatics, State Key Laboratory of Vascular Homeostasis and Remodeling, School of Basic Medical Sciences, Peking University, 38 Xueyuan Rd, Beijing, 100191, China; Department of Cardiology and Institute of Vascular Medicine, State Key Laboratory of Vascular Homeostasis and Remodeling, Peking University Third Hospital, 49 Huayuanbei Road, Beijing 100191, China; School of Sports Medicine, Wuhan Sports University, No. 461 Luoyu Rd. Wuchang District, Wuhan 430079, Hubei Province, China; National Clinical Research Center for Cardiovascular Diseases, State Key Laboratory of Cardiovascular Disease, Fuwai Hospital, National Center for Cardiovascular Diseases, Chinese Academy of Medical Sciences and Peking Union Medical College, Beijing, China

## Abstract

Coronary artery disease (CAD) is a complex, multifactorial, and serious condition influenced by genetic, environmental, and lifestyle factors. Thus, it is crucial to develop strategies to predict the risk of CAD for individuals. Plasma proteomics provides a powerful framework for identifying novel biomarkers, discovering potential therapeutic targets, and further improving risk stratification. Here, we examined the association between 2,919 plasma proteins and incident CAD in the UK Biobank cohort (n=35,778). As a result, we identified 576 proteins significantly associated with CAD and found significant alterations in key biological pathways, including signal transduction, immune regulation, and chemotaxis, before CAD onset. Subsequently, we developed machine learning models to predict CAD onset at different time intervals (5 years, 10 years, over 10 years, and entire cohort), demonstrating superior performance over models based on polygenic risk scores (ΔAUC = 0.052), and Pooled Cohort Equations (ΔAUC = 0.049). Notably, the integration of PRS with proteomic data resulted in a marked enhancement in predictive accuracy (AUC = 0.779), comparable to the full model (AUC = 0.780). Key plasma protein predictors, including MMP12, GDF15, and EDA2R, showed sustained importance across models predicting CAD onset at multiple time points. Additionally, Mendelian randomization analysis provided robust evidence for a causal relationship between six plasma proteins and CAD, including MMP12, LPA and PLA2G7, highlighting their potential as therapeutic targets. In conclusion, our study elucidates the plasma proteome associated with CAD, reveals underlying pathogenic mechanisms, and provides valuable insights for identifying high-risk individuals and advancing precision medicine.

## Introduction

Coronary artery disease (CAD) is one of the leading causes of morbidity and mortality worldwide[1]. As such, early assessment of individual CAD risk is crucial for the effective implementation of primary prevention strategies[2]. Several risk prediction models have been developed to assess CAD risk in asymptomatic individuals, including the widely used Pooled Cohort Equations (PCE), which is primarily employed to estimate the 10-year incidence of atherosclerotic cardiovascular disease events in primary prevention populations[3–7]. However, comprehensive risk assessment for CAD typically requires the integration of predictive factors from multiple domains[8, 9]. This process is often time-consuming and costly, severely limiting its clinical applicability. Consequently, the development of novel, precise predictive factors for CAD risk is paramount to advancing the field of precision medicine.

The implication of precision medicine currently utilizes polygenic risk scores (PRS) derived from disease susceptibility loci to assess disease risk, marking significant progress[10–12]. However, the intricate processes of gene transcription and translation into functional proteins obscure potential disease-associated genetic variants, thus impeding our understanding of the genomic-disease link and restricting the discovery of targeted therapies[13]. Circulating proteins provide a comprehensive reflection of an individual’s health status, integrating genetic, lifestyle, environmental, comorbidity, and medication effects[14, 15], presenting a promising opportunity for precision medicine. Currently, several plasma proteins have been identified to be potentially associated with the development of CAD[16–19], Nevertheless, these studies are often constrained by cohort size and the number of protein measurements, limiting their broader applicability. Moreover, several proteomics-based predictive studies employ cross-sectional designs[20, 21], thereby neglecting the causal relationship between plasma proteins and disease, and failing to capture the proteomic alterations that precede disease onset.

Recently, the UK Biobank (UKB) released plasma proteomic data from over 50,000 individuals, covering approximately 3,000 proteins, offering a unique opportunity to leverage large-scale longitudinal proteomic cohorts for disease prediction[22–24]. Here, using Cox proportional hazards (PH) models, we first identified the plasma proteins associated with CAD risk and explored key biological pathways altered before disease onset. And then these CAD-associated proteins were utilized to develop machine learning-based risk prediction models, assessing their predictive accuracy across the entire cohort as well as at specific time points (including 5, 10, and over 10 years), while also identifying plasma proteins with strong predictive power. Furthermore, we compared the performance of the proteomic model with PRS, PCE scores, and their combinations. Finally, Mendelian randomization analysis was applied to explore the genetic basis linking plasma proteins with CAD.

## Methods

### UKB participants

UKB is a large-scale, prospective cohort study that serves as a comprehensive global biomedical database, containing extensive genetic, proteomic and phenotypic data. Participants were recruited between 2006 and 2010 from 22 centers across the UK, encompassing 502,156 individuals aged 40-69 years at baseline. Considering the differences in proteomic characteristics and disease susceptibility across ancestries[25], this study selected the European participants in the UKB Pharma Plasma Proteome (UKB-PPP) for analysis. Peddy, a machine learning-based software tool that predicts sample ancestry using genetic loci[26], was employed to identify European populations, defined as those with Pr(EUR) > 0.98[23]. Moreover, participants with a history of CAD at baseline, those with more than 20% missing proteomic data, and individuals lacking PRS were excluded from the analysis. The detailed sample inclusion criteria are illustrated in **Supplementary Fig. S1**. Ultimately, a total of 35,778 participants were included in the analysis, with a median follow-up of 14.5 years, during which 1,458 participants experienced CAD events. This study utilized data from the UK Biobank Resource under approved application number 87841.

### Proteomic measurement and processing

UKB-PPP conducted proteomic profiling using EDTA plasma samples from approximately 54,000 participants[27]. Details regarding sample selection, processing and quality control are described in previous publications[22]. In short, UKB-PPP employed the Olink Explore 3072 PEA platform to measure 2,923 unique proteins across four Olink panels (cardiovascular metabolism, inflammation, neurology, and oncology). The study cohort includes a random subset of 46,595 participants, 6,356 participants selected by UKB-PPP, and 1,268 participants who took part in repeated imaging studies during the COVID-19 follow-up phase[22]. We removed proteins with missingness samples rates greater than 20% (CTSS, GLIPR1, NPM1, PCOLCE). Subsequently, missing protein measurements were imputed using the *k*-nearest neighbor (KNN) (*k* = 10) imputation method. Before further analysis, each imputed protein level was inverse rank normalized and standardized to have a mean of 0 and a standard deviation of 1[28]. The final plasma protein dataset included 35,778 individuals and 2,919 unique proteins.

### Clinical risk information

The clinical variables of the study cohort are summarized in **Table 1**, including age, sex, Townsend Deprivation Index (TDI), body mass index (BMI), systolic blood pressure (SBP), smoking status, total cholesterol, high-density lipoprotein cholesterol (HDL-C) levels, as well as blood pressure-lowering medications and history of diabetes. The definitions of history of antihypertensive medication and history of diabetes followed previous studies[29]. A detailed list of UKB fields used is provided in **Supplementary Table S1**. For missing data, categorical variables were imputed using the mode, and then continuous variables were imputed using the KNN (*k* = 10) method. In addition, The R package PooledCohort was used to calculate the PCE scores.

**Table 1.**
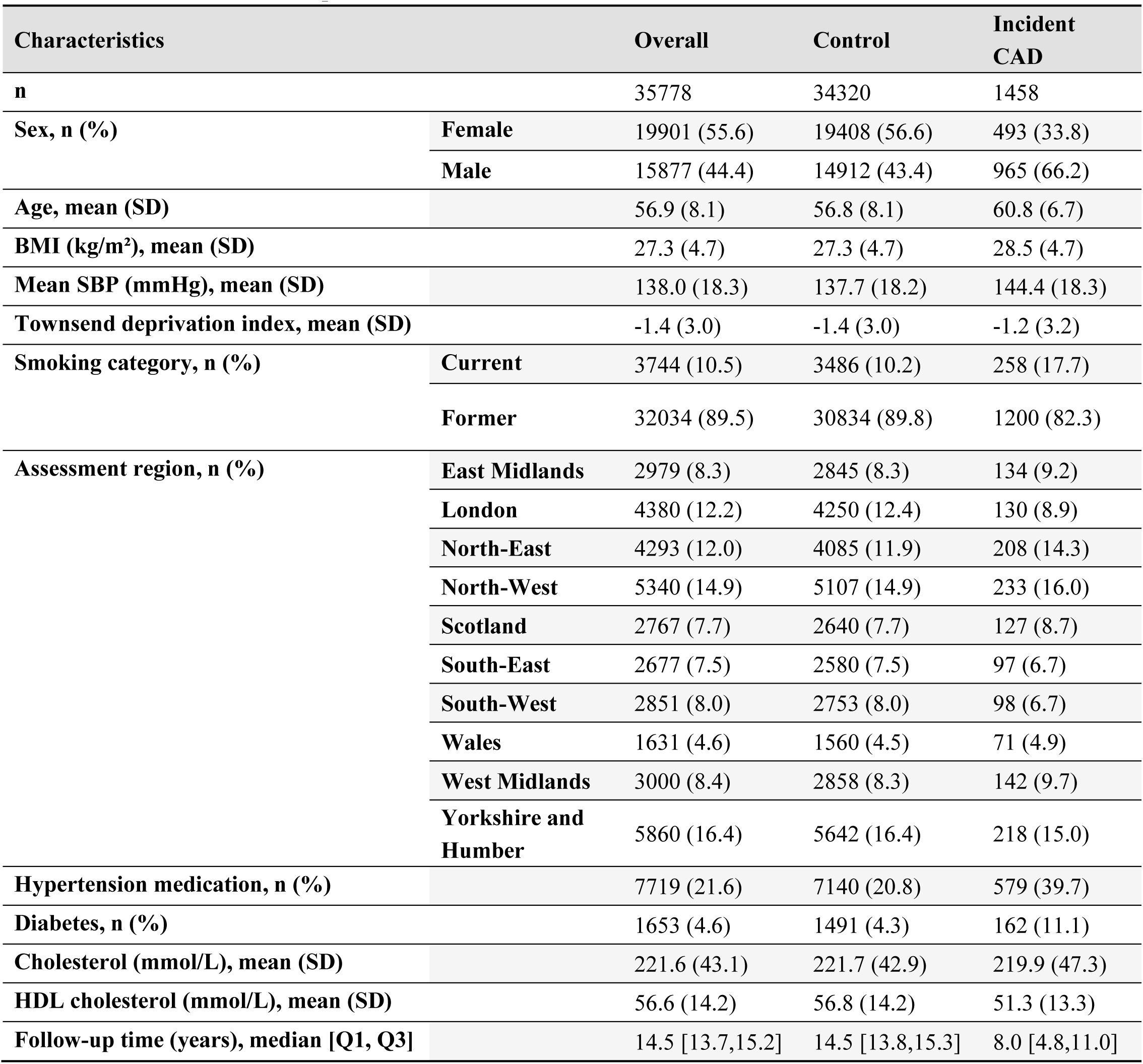
The descriptive characteristics at baseline.

### Incident CAD definitions

The UKB data is linked with hospital episode statistics, which records diagnostic information using the International Classification of Diseases versions 9 (ICD-9) and 10 (ICD-10), while surgical procedures are coded using Office of Population Censuses and Surveys: Classification of Interventions and Procedures, Fourth Edition (OPCS-4) [30]. To determine participants diagnosed with CAD events, our study adopted the definition outlined in previous reports [29, 31]. Specifically, we extracted the CAD-related events from several fields, comprising ICD-9, ICD-10, OPCS-4, noncancer illness code, operation code, the vascular/heart problems, as well as primary and secondary causes of death recorded with ICD10 codes. **Supplementary Table S2** presents a detailed list of the fields used to define CAD.

Incident CAD was defined as the diagnosis of any CAD event occurring more than six months after the baseline UKB assessment. The time of incident CAD was ascertained in accordance with the earliest onset date documented in the relevant UKB fields. To illustrate, for participants with multiple CAD event records, only the earliest recorded incident of CAD was preserved. The follow-up time was calculated as the number of days from the assessment date to the occurrence of the event of interest, competing death events from other causes, or the date of the last recorded event (September 13, 2023).

### Mendelian randomization analysis

The protein quantitative trait loci (pQTL) data for European ancestry were obtained from the UKB-PPP resource (https://metabolomips.org/ukbbpgwas/)[22]. Details of the pQTL analysis are described in previous publications[22]. The GWAS summary statistics for CAD were downloaded from the IEU database (ieu-a-7)[32]. Single nucleotide polymorphisms (SNPs) with p-values less than 5 × 10^−8^ were selected as instrumental variables. Clumping was applied to remove SNPs in linkage disequilibrium, and the F-statistic for each SNP was calculated to exclude weak instruments (F-statistic < 10). Data harmonization was performed to ensure consistency between the exposure and outcome. Mendelian randomization (MR) analysis was then conducted using five distinct methods, including inverse variance weighting (IVW), MR-Egger, and weighted median, weighted mode and simple mode. Sensitivity analyses, such as heterogeneity tests and horizontal pleiotropy tests, were conducted to assess the robustness of the results. All analyses were performed using the R package “TwoSampleMR“[33].

### Statistical analyses

The Cox PH model and the machine learning-based predictive model were developed and evaluated using ten-fold cross-validation. Specifically, the study cohort was partitioned into ten folds according to the geographic locations of the 22 assessment centers (**Supplementary Table S3**). Nine of the ten folds were employed for model training, while the remaining fold functioned as the test set. This procedure was iteratively repeated until each fold had been utilized for both training and validation. Hyperparameter optimization for the machine learning model was performed within the training set using five-fold cross-validation on randomly partitioned subsets.

Cox PH models were applied to examine the association between standardized plasma protein expression levels and the incidence of CAD. Model 1 adjusted for age and sex, while Model 2 further accounted for additional covariates, including the TDI, BMI, SBP, smoking status, antihypertensive medication use, history of diabetes, and concentrations of total cholesterol and HDL-C. Proteins with a Benjamini-Hochberg (BH) adjusted P-value < 0.05 were considered significantly associated with incident CAD. Enrichment analysis was performed using the clusterProfiler package, with CAD-related plasma proteins as input and 2,919 proteins as the background genes.

LightGBM (LGBM) is a machine learning algorithm built on gradient boosting decision trees, renowned for its robust performance and widespread application in clinical risk prediction[34–36]. In this study, we employed LGBM to predict incident CAD, with hyperparameter optimization conducted using Optuna[37], aiming to maximize the area under the curve (AUC) on the test set. Upon determining the optimal hyperparameters, protein importance was assessed using the training set, with proteins ranked according to their mean importance values derived from ten-fold cross-validation. A forward stepwise strategy was then implemented, in which proteins were progressively incorporated into the LGBM model according to their importance rankings, followed by performance evaluation on the test sets. Shapley Additive Explanations (SHAP) values were employed to elucidate the contribution of each protein to the predictive model. The disparities between the AUCs of the two models of interest were evaluated using the DeLong test.

Data visualization and analysis in this study were conducted using R (v.4.3.1) and Python (v.3.12.4). The following libraries were employed: clusterProfiler (v.4.10.1), lifelines (v.0.29.0), MLstatkit (v.0.1.7), Optuna (v.4.0.0), PooledCohort (v.0.0.2), LightGBM (v.4.5.0), scikit-learn (v.1.5.1), SHAP (v.0.46.0), and TwoSampleMR (v.0.6.8).

## Results

### Overview of cohort characteristics

We conducted a cohort study comprising 35,778 participants of European ancestry from UKB, all of whom were free of CAD at baseline. Baseline characteristics of the cohort are summarized in **Table 1**. The cohort’s mean age was 56.9 years (Standard Deviation [SD] 8.1), with females comprising 55.6% of the population. During a median follow-up of 14.5 years (interquartile range [IQR] 13.7-15.2), a total of 1,458 individuals (4.08%) were diagnosed with CAD.

### Associations between proteins and incident CAD

Subsequent to protein data quality control, a total of 2,919 proteins were preserved for the examination of their association with CAD. In model 1, 1488 proteins exhibited significant association with CAD, adjusting for sex and age (**Fig. 2A and Supplementary Table S4**). To further mitigate potential confounding factors, model 2 additionally considered adjustments for the TDI, BMI, SBP, smoking status, blood pressure-lowering medications, history of diabetes, as well as total cholesterol and HDL-C levels (**Fig. 2B and Supplementary Table S4**). In model 2, 593 proteins were found to be statistically significant following BH correction, of which 577 were consistently identified in model 1. MMP12, GDF15 and TNFrsF10b exhibited the most pronounced associations across both models. Specifically, MMP12 demonstrated a hazard ratio (HR) of 1.64 (P-adjust = 2.28 × 10^−61^) in model 1 and 1.45 (P-adjust = 3.06 × 10^−29^) in model 2. Likewise, GDF15 displayed an HR of 1.70 (P-adjust = 2.10 × 10^−58^) in model 1 and 1.44 (P-adjust = 2.52 × 10^−20^) in model 2. Aligned with these findings, GDF15 has garnered extensive recognition for its pivotal role in clinical research and translational advancements[38, 39].

**Fig. 1.**
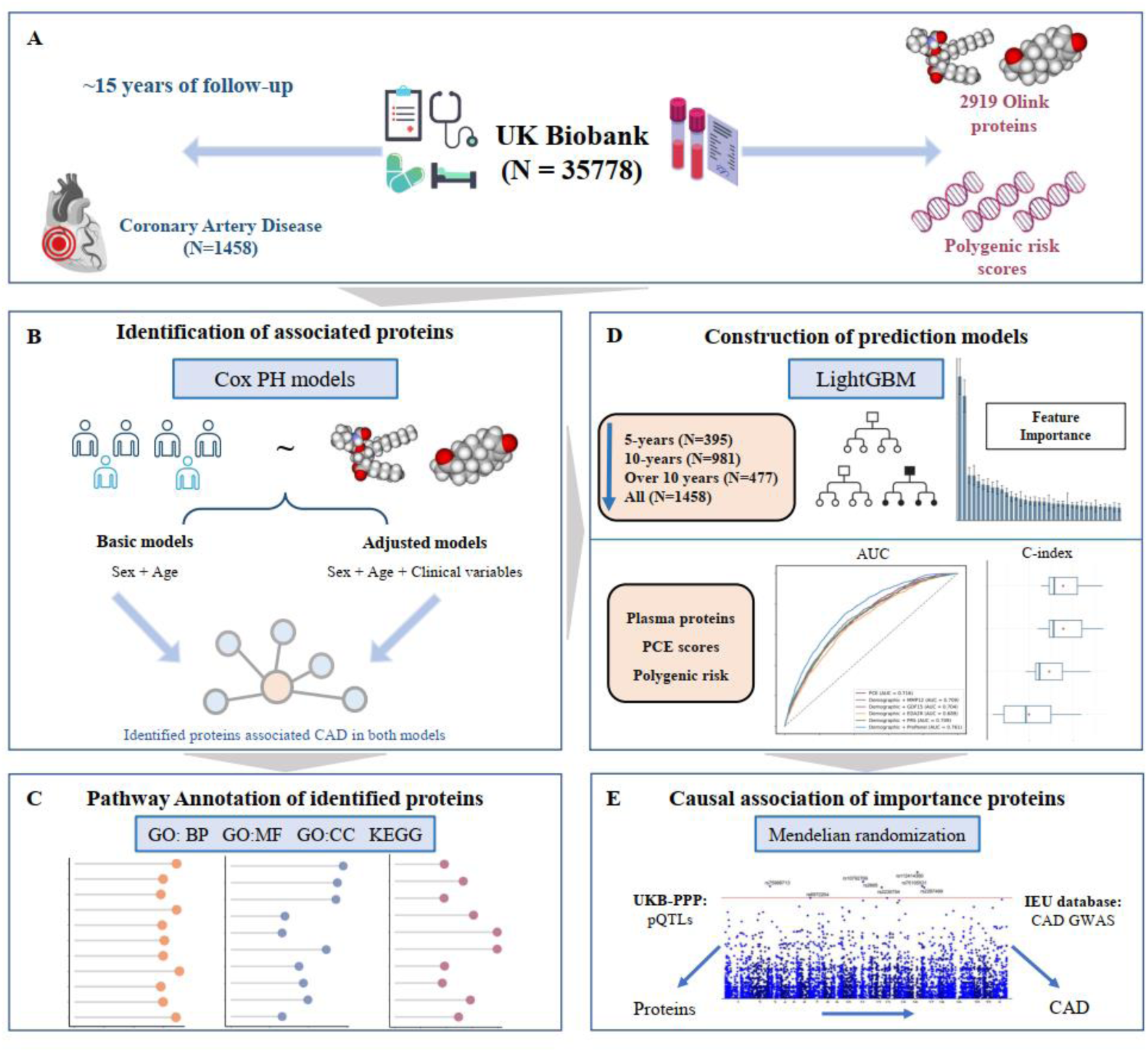
Study Overview. This study leverages the extensive plasma proteomic cohort from the UK Biobank, with a median follow-up of 14.5 years, to construct predictive models for coronary artery disease (CAD) (**A**). Cox proportional hazards (PH) models were employed to evaluate associations between individual plasma proteins and CAD (**B**), and significant proteins were further analyzed to illustrate alterations in biological pathways preceding CAD onset (**C**). Additionally, we employed the LightGBM machine learning model to evaluate the predictive value of plasma proteins by analyzing feature importance and SHAP values across specific time intervals, including 5 years, 10 years, over 10 years, and the entire cohort. The CAD prediction models were subsequently constructed within a 10-fold cross-validation framework, with their performance meticulously compared to models based on demographics, polygenic risk scores (PRS), Pooled Cohort Equations (PCE), and their various combinations (**D**). Finally, publicly available genetic datasets, including protein quantitative trait loci (pQTLs) from the UKB-PPP and summary-level genome-wide association study (GWAS) data for CAD from IEU, were utilized. Mendelian randomization analysis was conducted to identify key proteins with causal links to CAD, providing insights into potential therapeutic targets (**E**).

**Fig. 2.**
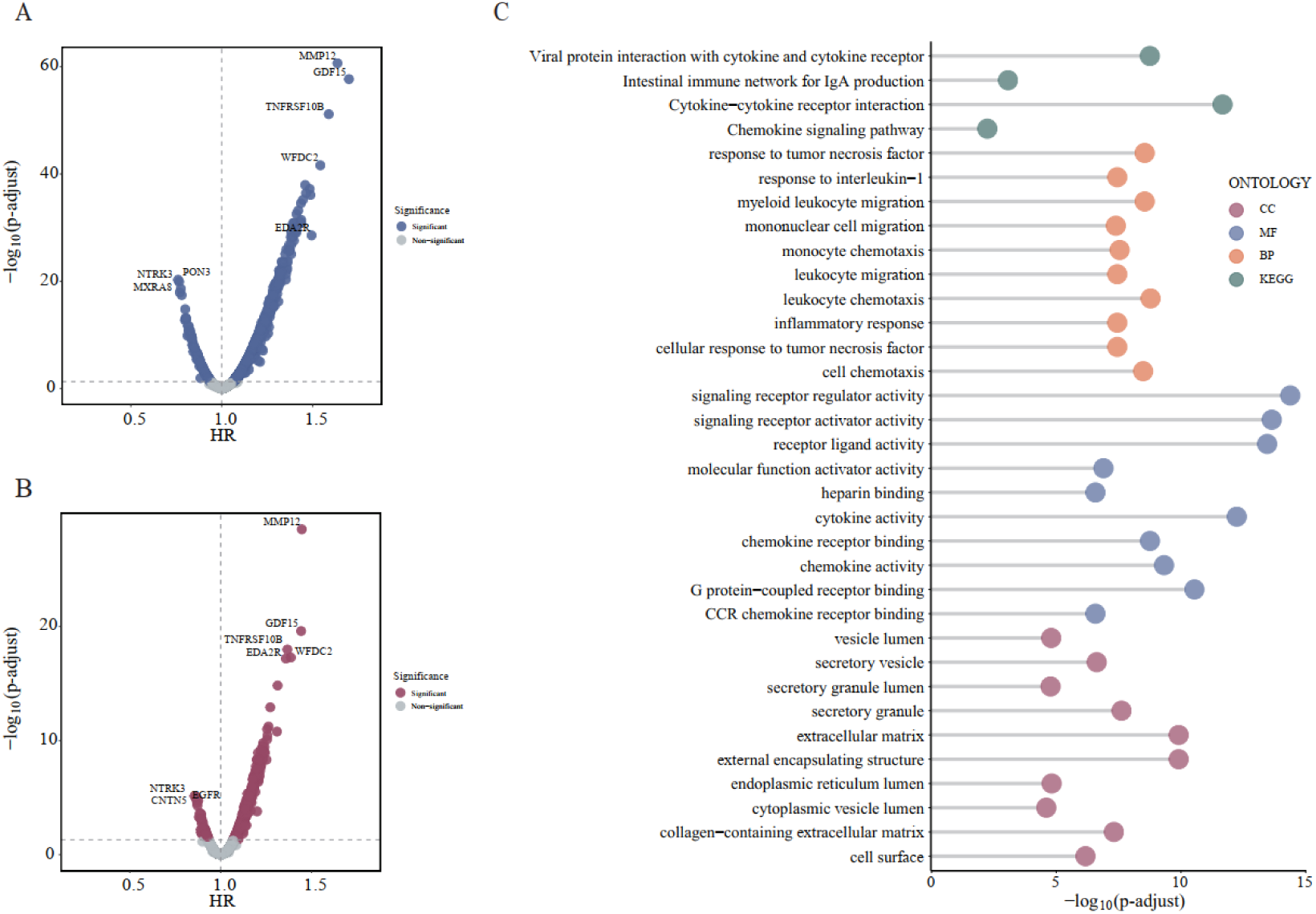
Proteome-wide associations with incident CAD. (**A**) Proteins significantly associated with CAD in Model 1 after adjustment for sex and age. The x-axis represents the relative risk (hazard ratio, HR), with blue dots denoting proteins with an adjusted P-value < 0.05 after Benjamini-Hochberg correction. (**B**) Proteins significantly associated with CAD in Model 2 after additional adjustments for sex, age, Townsend Deprivation Index (TDI), body mass index (BMI), systolic blood pressure (SBP), smoking status, antihypertensive medication use, diabetes history, as well as total cholesterol and high-density lipoprotein cholesterol (HDL-C) levels. Red dots indicate proteins with an adjusted P-value < 0.05 after Benjamini-Hochberg correction. (**C**) Enrichment analysis of proteins significantly associated with CAD in both Model 1 and Model 2. Different colors represent Gene Ontology (GO) terms for molecular function (MF), cellular component (CC), biological process (BP), and Kyoto Encyclopedia of Genes and Genomes (KEGG) pathways, showcasing the top ten significant results for each ontology.

To deepen our understanding of how the identified proteins contribute to the pathogenesis of CAD. Functional enrichment analysis was implemented on significant proteins, several pathways associated with the regulation of signal transduction and immune response are profoundly enriched (**Fig. 2C and Supplementary Table S5**). Of particular note, chemotaxis-related pathways are consistently identified across multiple ontologies, including biological processes, molecular functions, and KEGG pathways, suggesting that chemotaxis may play a critical role in the pathogenesis of CAD.

### Construction of CAD prediction models

The proteins consistently identified in both models were employed to construct machine learning-based predictive models for incident CAD. Proteins were ranked according to their feature importance in the predictive task, with MMP12, GDF15, EDA2R, CHCHD10 and HAVCR1 emerging as the top five predictors (**Fig. 3A and Supplementary Table S6**). Considering clinical applicability, we implemented two strategies to evaluate the AUC of the model with the aim of selecting an optimal protein panel. In the first strategy, we began by constructing a model using the protein ranked highest in importance, then iteratively refined the model by incorporating additional proteins based on their feature importance, while assessing the AUC at each stage (**Supplementary Table S7**). The second strategy recognized that demographic factors such as sex and age are readily available and could potentially enhance the model’s predictive accuracy. Thus, we incorporated sex and age into the initial model and repeated the refinement process as described above (**Supplementary Table S8**). DeLong test was utilized to evaluate the impact on AUC before and after each model update. With the gradual inclusion of the first 12 proteins (up to PZP), the average AUC of models constructed using both strategies showed a significant improvement (**Fig. 3B**). Initially, the protein-only model showed a lower AUC than the model with demographic factors, but the performance gap attributed to sex and age gradually narrowed as more proteins were included. Both models stabilized at an AUC of approximately 0.76 after incorporating the first 29 proteins (up to ICOSLG), which were selected as the final proteome for constructing the predictive model (ProPanel).

**Fig. 3.**
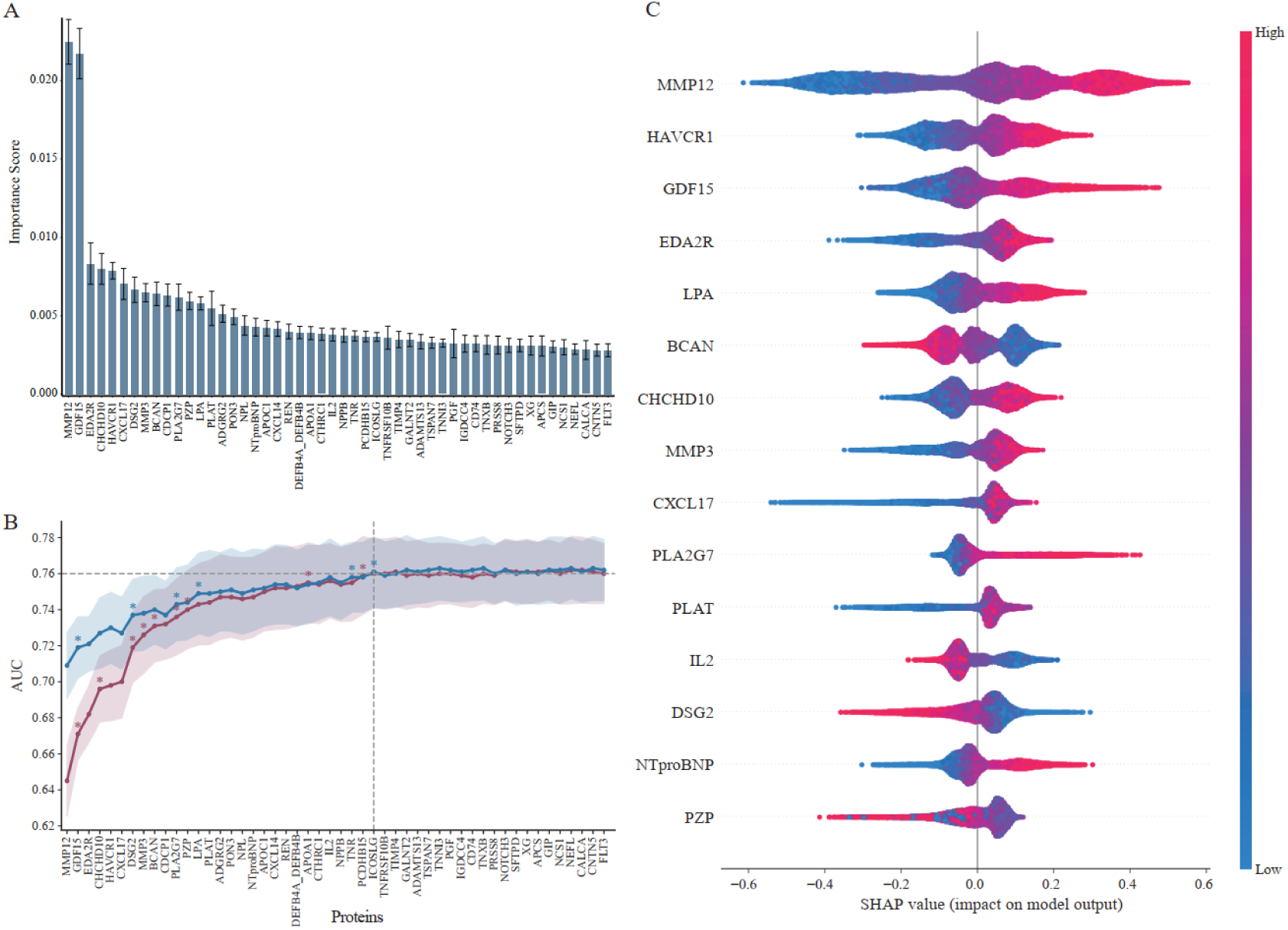
Contributions of plasma proteins to CAD prediction models. (**A**) Standardized feature importance of proteins, with error bars derived from cross-validation results. (**B**) The average area under the curve (AUC) derived from ten-fold cross-validation, before and after model updates, is shown. The red line represents the performance of the protein-only model, while the blue line corresponds to the model that includes sex and age features from the outset. The shaded area indicates the standard error obtained from cross-validation. An asterisk (*) denotes a statistically significant improvement in AUC following the addition of a new protein (P < 0.05, DeLong test). (**C**) Shapley additive explanations (SHAP) values were used to illustrate the contribution of important proteins to the predictive model. The range occupied by proteins on the x-axis reflects their contribution to CAD prediction, with a larger range indicating a greater contribution to the model. The color of the points represents the expression level of plasma proteins, with blue to red indicating an increasing expression level. Points located on the right side of the y-axis are associated with a higher likelihood of CAD occurrence, while those on the left side indicate a tendency towards health.

We utilized SHAP to analyze the top 15 protein to assess the contributions of proteins to the CAD prediction model (**Fig. 3C**). MMP12 exhibited the widest range of SHAP values. Elevated expression of MMP12 was strongly associated with an increased risk of CAD, whereas lower expression levels were correlated with a healthier phenotype, underscoring its crucial role in CAD prediction. In contrast, several proteins, such as BCAN, IL2, and DSG2, displayed a protective effect against CAD when present at higher plasma concentrations.

We further investigated the predictive potential of plasma proteins for CAD across varying disease onset timelines. Participants were stratified into three cohorts based on the time to onset: 5 years, 10 years, and over 10 years. For each subgroup, we independently analyzed feature importance (**Supplementary Table S6, S7, S8 and Fig. S2**). As a results, the model predicting CAD incidence within 5 years demonstrated the highest predictive performance (AUC = 0.768), followed by the 10-year model (**Supplementary Fig. S3**). Notably, proteins MMP12 and GDF15, identified as pivotal across multiple time horizons, serve as critical factors in disease risk stratification. Remarkably, the declining predictive performance for CAD onset beyond 10 years suggests that long-term risk may be influenced by additional factors not fully captured by the analyzed proteomic signatures. Together, these findings emphasize the temporal dynamics of CAD risk prediction and the critical role of specific plasma proteins in guiding early intervention strategies.

### Protein models compared with clinical predictors and polygenic risk scores

By modeling the risk for incident CAD, we evaluated the predictive power of PCE scores and demographic factors (sex and age) combined with individual proteins, PRS, ProPanel, as well as their integrated combinations. To assess the AUC across different feature sets, we conducted ten-fold cross-validation on the entire cohort. The individual protein MMP12, when combined with demographic factors, demonstrated strong predictive power (AUC = 0.709), comparable to that of the demographic-PRS model (AUC = 0.709), though slightly inferior to the PCE-based model (AUC = 0.716) (**Fig. 4A**). When combined with demographics, ProPanel exhibited superior predictive performance, outperforming both PCE-based (ΔAUC = 0.045) and PRS-based models (ΔAUC = 0.052) (**Fig. 4A**). Furthermore, integrating ProPanel with demographics and PRS resulted in an additional AUC improvement, reaching 0.779, closely matching the predictive accuracy of the PCE-ProPanel-PRS model (AUC = 0.780), which requires supplementary clinical variables (**Fig. 4B**). Cohort analysis using the feature combination of ProPanel, demographic factors, and PRS also yielded robust performance, with a C-index of 0.789 (**Fig. 4C**). Here, MMP12, GDF15, and EDAR2 as indispensable biomarkers for CAD risk are clearly highlighted, with ProPanel showcasing remarkable predictive capability, further strengthened by the integration of PRS.

**Fig. 4.**
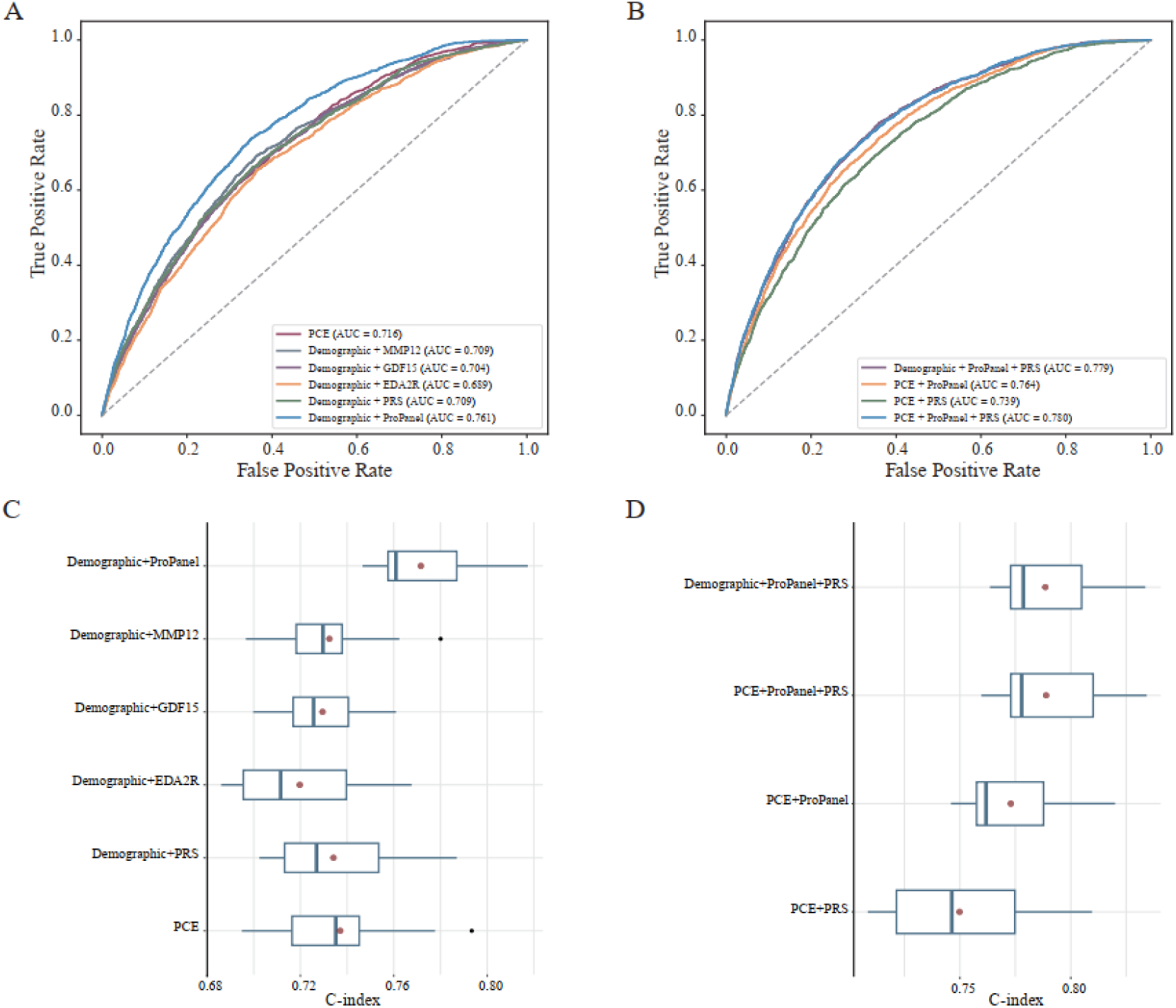
Performance of the CAD Prediction Models. (**A**) Evaluation of the predictive performance of PCE scores and demographic factors (sex and age) combined with MMP12, GDF15, EDA2R, PRS, and ProPanel. The ProPanel model incorporates the 29 most predictive proteins. PCE scores were derived from a clinical variable-based model including TDI, BMI, SBP, smoking status, antihypertensive medications, diabetes history, and levels of total cholesterol and HDL-C. The average AUC from ten-fold cross-validation is presented. (**B**) Comparison of AUC values for combinations of ProPanel, PRS, and PCE scores. The average AUC from ten-fold cross-validation is presented. (**C**) Performance analysis of PCE scores and demographic factors combined with MMP12, GDF15, EDA2R, PRS, and ProPanel using the Cox PH model. Red dots represent the average C-index derived from 10-fold cross-validation. (**D**) Assessment of prediction accuracy for combinations of ProPanel, PRS, and PCE scores within the Cox PH model.

**Fig 5.**
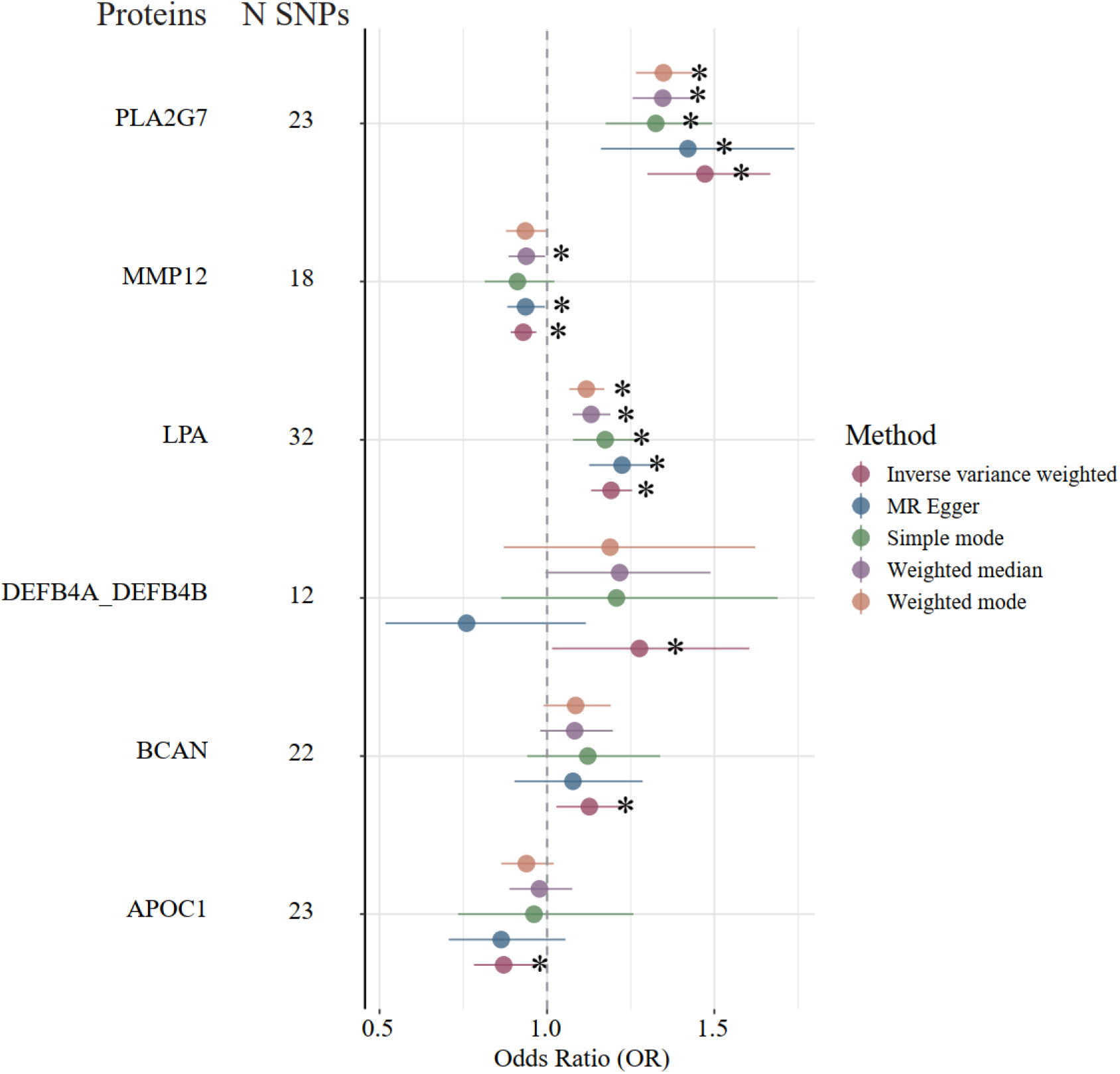
The forest plot illustrates plasma proteins significantly associated with CAD, as identified through Mendelian randomization analyses using the inverse variance weighting (IVW) method (P-value < 0.05). Distinct colors denote the five analytical methods applied: IVW, MR-Egger, weighted median, weighted mode, and simple mode. An asterisk (*) indicates that the corresponding protein achieved a P-value below 0.05 within a specific method. The horizontal lines denote the 95% confidence intervals, while “N SNPs” indicates the number of instrumental variables used in each analysis.

### Causal effects between plasma proteins and CAD

Proteins may either play a causal role in the disease process or emerge as consequences of the disease. Leveraging publicly available genetic resources, such as proteomics-based pQTLs and summary-level GWAS data for CAD, enables the identification of potential causal proteins. From the top 30 most important proteins, Mendelian randomization analysis identified six with significant causal associations to incident CAD (IVW, P-value < 0.05). The top three were MMP12 (β = −0.0735, P-value = 4.36 × 10^−4^, OR = 0.929, 95% CI: 0.892–0.968), PLA2G7 (β = 0.386, P-value = 1.16 × 10^−9^, OR = 1.472, 95% CI: 1.300–1.667), and LPA (β = 0.175, P = 2.27 × 10^−11^, OR = 1.191, 95% CI: 1.132–1.254). Heterogeneity tests showed no significant heterogeneity for MMP12 (IVW: Q = 15.77, P-value = 0.540; MR-Egger: Q = 15.67, P-value = 0.477). Egger intercept tests revealed no directional pleiotropy for MMP12 (intercept = −0.0017, P-value = 0.747), PLA2G7 (intercept = 0.0048, P-value = 0.665), or LPA (intercept = −0.0097, P-value = 0.425). Key proteins with a causal role in CAD pathogenesis emerge as promising therapeutic targets, warranting further investigation to clarify their role in prevention and treatment.

## Discussion

Plasma proteomics offers valuable opportunities for disease risk prediction and novel therapeutic target discovery. In this study, we conducted an association analysis between CAD and plasma proteomics using data from the UKB-PPP European cohort, uncovering a comprehensive array of plasma proteins intricately linked to CAD risk. These CAD-related proteins were markedly enriched in pathways linked to immune response and signal transduction. We then developed plasma protein-based models for CAD risk prediction across the entire cohort, as well as at specific time points (including 5, 10, and over 10 years). Key proteins, such as MMP12, GDF15, EDA2R, CHCHD10, and HAVCR1, were identified as pivotal contributors to the model’s enhanced predictive accuracy. The plasma protein-based model demonstrated remarkable superiority over models based on polygenic risk scores (PRS) and Pooled Cohort Equations (PCE) in terms of predictive performance. Moreover, the combination of plasma proteins and PRS achieved excellent predictive accuracy, comparable to the full model. Mendelian randomization analysis reinforced the causal relationship between six plasma proteins, particularly MMP12, LPA, and PLA2G7, stressing their potential as predictive biomarkers and therapeutic targets for CAD.

We identified plasma proteins associated with CAD that were significantly enriched in chemotactic pathways, including CCR chemokine receptor binding and chemokine activity. Chemokines are signaling molecules that mediate immune cell recruitment, regulate cell homeostasis, and activate various immune cell types and subpopulations[40]. The production of chemokines and the activation of their receptors establish a positive feedback loop, recruiting monocytes, neutrophils, and lymphocytes to atherosclerotic plaques[40]. This process underscores the integral role of chemokines in the progression of cardiovascular diseases. Additionally, extensive epidemiological studies unveil the crucial role of chemokines and their receptors in a range of cardiovascular diseases, including atherosclerosis, myocardial infarction, and heart failure[41]. For instance, Georgakis et al. demonstrated that elevated circulating levels of CCL2 are linked to an increased risk of long-term cardiovascular mortality[42]. Despite these findings, the exact molecular mechanisms by which chemokines contribute to cardiovascular pathology remain incompletely understood, further investigation is needed to elucidate their precise roles.

Our study further validated several plasma proteins previously identified in the literature, reinforcing their relevance to CAD pathogenesis. Notably, we observed that elevated GDF15 expression is strongly associated with an increased risk of CAD, consistent with findings by Hagström et al., who demonstrated its prognostic value in cardiovascular outcomes [43], Additionally, Takaoka et al. provided experimental evidence that blocking GDF15 in murine models not only mitigated cachexia but also decelerated the progression of heart failure[44]. Another key protein, lipoprotein(a) (Lp[a]) (LPA), emerged as a robust risk factor for cardiovascular diseases, corroborating its widely recognized role as a pathogenic determinant. Numerous studies have established LPA’s contribution to atherogenesis, thrombosis, and inflammation[45–47], further emphasizing its clinical significance in cardiovascular risk stratification and therapeutic targeting. Interestingly, our cohort study and Mendelian randomization analysis revealed a significant association between platelet-activating factor acetylhydrolase (PLA2G7) and CAD. Mutations in the gene encoding this protein are linked to Lp-PLA2 activity[48]. However, a meta-analysis conducted a decade ago concluded that, while PLA2G7 variants influence Lp-PLA2 activity, these variants are not associated with cardiovascular risk markers, coronary atherosclerosis, or CAD[49]. By leveraging a substantially larger cohort, our study challenges this earlier conclusion, presenting compelling evidence to support the potential of PLA2G7 as a biomarker for CAD.

MMP12 emerged as a particularly noteworthy protein in our study, exhibiting strong predictive power for CAD risk and a robust causal relationship as supported by Mendelian randomization analysis, alongside confirmation through sensitivity analyses. Elevated baseline expression levels of MMP12 were associated with an increased risk of CAD, aligning with previous reports[50, 51]. Interestingly, however, Mendelian randomization revealed a protective causal effect of MMP12 on CAD, a discrepancy that mirrors observations by Henry et al. in their study on heart failure[52]. We hypothesize that this apparent inconsistency may stem from the cross-sectional nature of both pQTL studies and disease GWAS, where CAD development could potentially downregulate the expression of certain proteins, including MMP12. This accentuates the need for longitudinal cohort studies to capture dynamic changes over time, offering greater clarity on the temporal and causal relationships between protein expression and CAD progression. Connecting the underlying mechanisms that regulate MMP12 expression and its role in disease pathophysiology, which could provide valuable insights into the intricate biological processes driving CAD.

This study has several limitations. Firstly, numerous genetic susceptibility loci for diseases exhibit significant racial heterogeneity. For instance, Ding et al. identified several Alzheimer’s disease susceptibility loci that show ancestry-specific variations across different populations[53]. Additionally, loss-of-function mutations in PCSK9 were discovered using sequencing data from individuals of African ancestry[54], suggesting the need to consider genetic diversity when assessing disease susceptibility. In this study, we developed a CAD prediction model using a European cohort, and its accuracy in other ethnic groups still needs to be assessed. Developing ethnicity-specific CAD prediction models and evaluating the effectiveness of drug targets tailored to different populations are crucial next steps. Secondly, while the CAD prediction model in this study underwent stringent data quality control and cross-validation, it was exclusively based on data from the UKB cohort. The lack of large-scale longitudinal proteomic datasets restricts the ability to independently validate and further evaluate the model’s performance in external cohorts.

Overall, this study offers a comprehensive investigation of CAD through a large-scale longitudinal proteomic cohort. Enrichment analysis revealed significant alterations in key pathways, such as signal transduction and immune modulation, occurring well before the clinical onset of the disease, with a particular emphasis on chemokines and their associated receptors. Additionally, by integrating these proteins into machine learning models, we achieved outstanding predictive performance, outperforming traditional models based on PRS and PCE scores. Finally, our study establishes causal relationships between several plasma proteins and CAD, providing valuable perspectives for the identification of therapeutic drug targets. In summary, our research offers vital insights for early intervention strategies in CAD and contributes to advancing the implementation of precision medicine.

## Supporting information

Supplemental table, will be used for the linked to the file on the preprint site

## Ethical approval

This study utilized data from the UK Biobank Resource under approved application number 87841.

## Data availability

The data used in this study are available from the UK Biobank under specific restrictions. As the data were accessed under license, they are not publicly available. Access to UK Biobank data can be requested through the standard application process (https://www.ukbiobank.ac.uk/registerapply/) under application ID 87841. All data supporting the findings of this study are included in the article and supplementary materials and can be obtained from the corresponding author upon request. Source data are provided with this article.

## Acknowledgements

This study was supported by the grants from the National Natural Science Foundation of China [62025102, 32301239, 82470373] and National High Level Hospital Clinical Research Funding (2023-GSP-ZD-2).

## Authors’ contributions

Q.-H.C. and C.L. conceived the project and thoroughly revised the manuscript. C.-H.Z., X.-W.J. and C.-M.C. collected, analyzed and interpreted the data, and drafted and revised the manuscript. X.-K.B. and G.-D.H. analyzed and interpreted the data. All authors read and approved the final manuscript.

## Supplementary Materials

**Fig S1.**
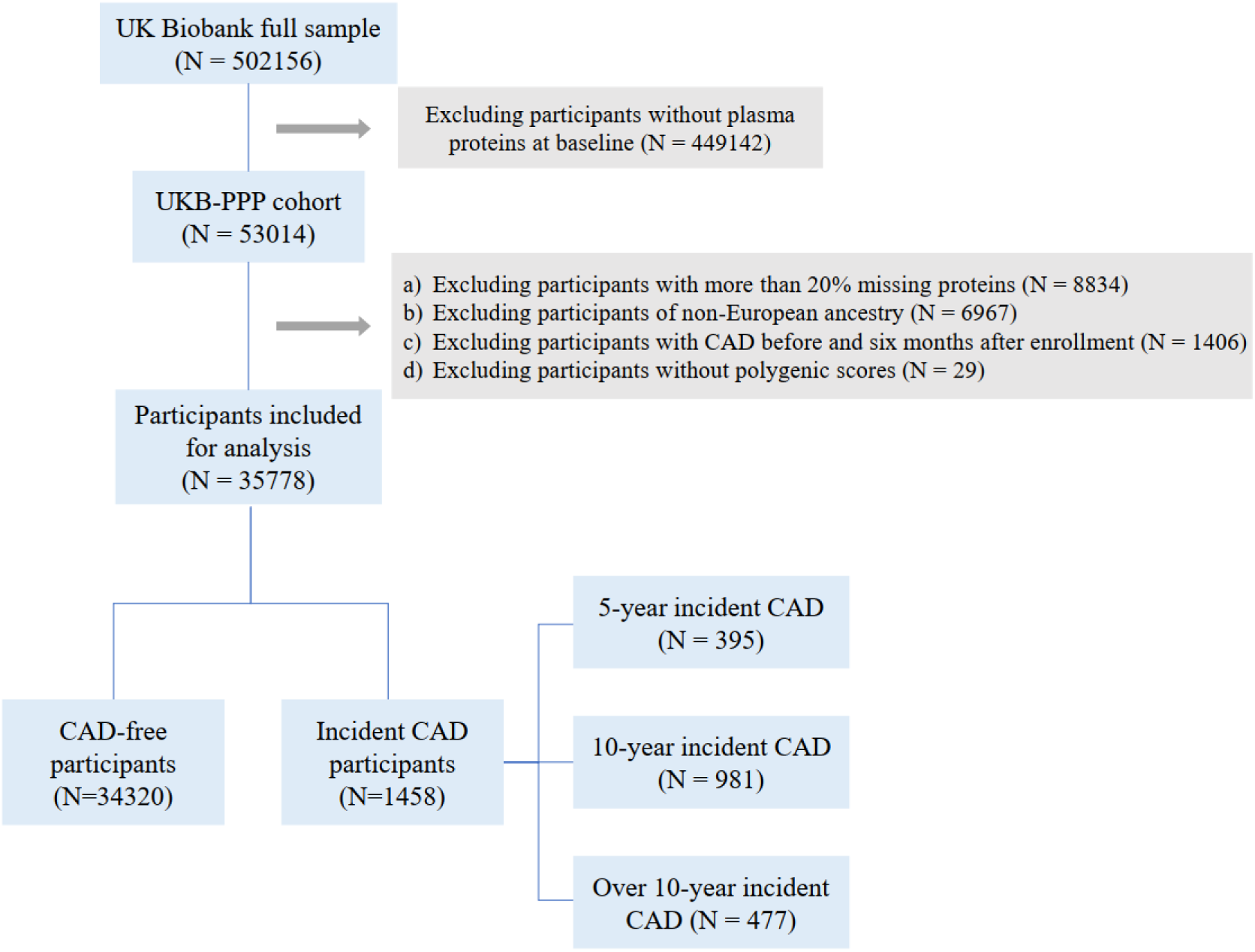
The participant inclusion process for this study. We excluded individuals from the UK Biobank cohort who did not undergo plasma proteomics analysis, were diagnosed with CAD within six months of recruitment, had more than 20% missing protein data, were of non-European ancestry, or lacked polygenic risk scores. The final cohort was further stratified into subgroups based on the time of CAD diagnosis: 5 years, 10 years, and over 10 years.

**Fig S2.**
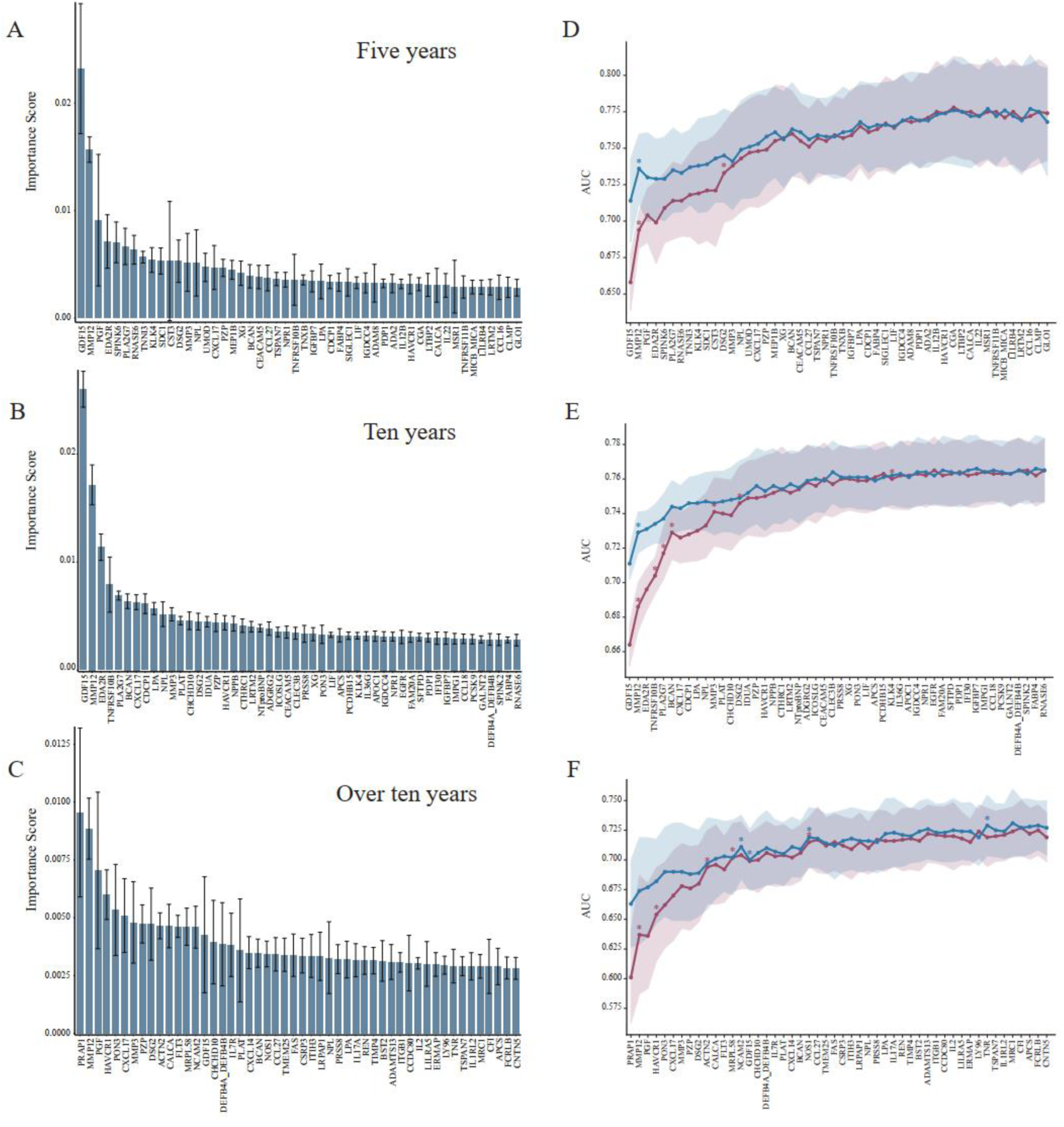
Construction of prediction models for different onset times. Standardized feature importance of proteins (A, B, C) and average area under the curve (AUC) from ten-fold cross-validation (D, E, F) for models constructed at different CAD onset times (5 years, 10 years, and over ten years). Error bars in (A, B, C) represent cross-validation results, while the shaded regions in (D, E, F) indicate the standard error. The red line reflects the performance of the protein-only model, whereas the blue line represents the model incorporating sex and age as baseline features. An asterisk (*) denotes a statistically significant improvement in AUC following the inclusion of a new protein (P < 0.05, DeLong test).

**Fig S3.**
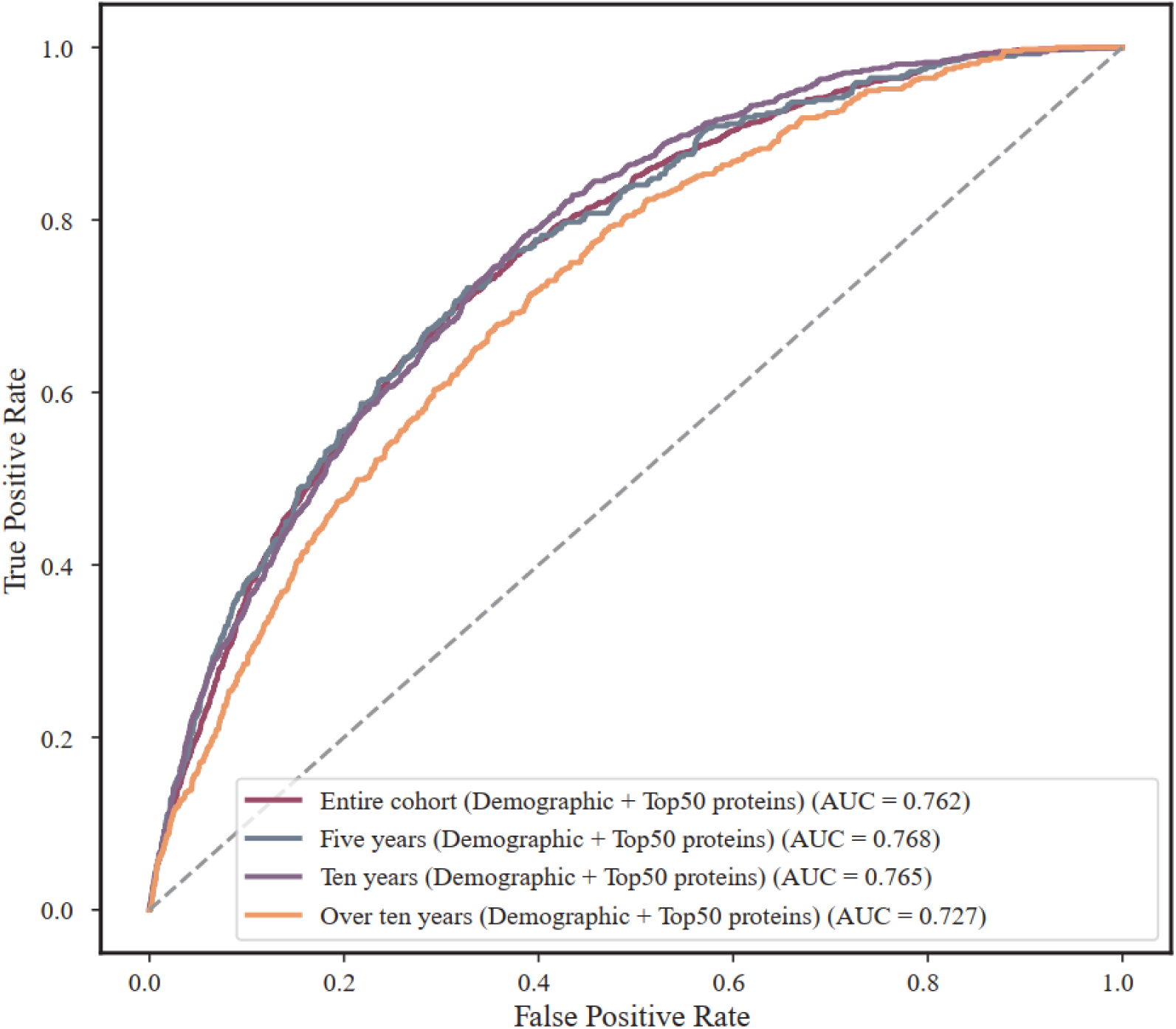
Performance of CAD prediction model with different onset time. The Area Under the Receiver Operating Characteristic Curve for models predicting CAD onset at the entire cohort, 5 years, 10 years, and over 10 years were all constructed using the top 50 most important proteins identified from each model, along with demographic factors. The average AUC from ten-fold cross-validation is presented..

**Table S1: The UKB Fields of clinical variables used in this study.**

**Table S2: Definition of Coronary Artery Disease (CAD).**

**Table S3: Geographic locations of the 22 assessment centers in UKB.**

**Table S4: The results of associations between plasma proteins and incident CAD.**

**Table S5: Enrichment results of proteins significantly associated in both models.**

**Table S6: Proteins ranked by feature importance in the predictive task.**

**Table S7: The performance of the model after adding proteins one by one.**

**Table S8: The performance of the model after adding proteins one by one (Include sex and age at the beginning of the model).**

**Table S9: The results of Mendelian two-sample randomization.**

